# Translating the Transcriptome: A Connectomics Approach for Gene-Network Mapping and Clinical Application

**DOI:** 10.1101/2025.08.08.25333301

**Authors:** Clemens Neudorfer, Bassam Al-Fatly, Barbara Hollunder, Ningfei Li, Garance M Meyer, Nanditha Rajamani, Konstantin Butenko, Matteo Vissani, Alan Bush, Nathaniel D Sisterson, Ehsan Tadayon, Frederic Schaper, Bahne Bahners, Lauren Hart, Savir Madan, Julianna Pijar, Philip Mosley, Harith Akram, Nicola Acevedo, David Castle, Susan Rossell, Peter Bosanac, Jill L Ostrem, Philip A Starr, Vincent JJ Odekerken, Rob MA deBie, Juan A Barcia, Himanshu Tyagi, Sameer A Sheth, Wayne K Goodman, Veerle Visser-Vandewalle, Martijn Figee, Darin D Dougherty, Ludvic Zrinzo, Eileen Joyce, Daniel Corp, Juho Joutsa, Thomas Picht, Katharina Faust, Andrea A Kühn, Christos Ganos, Jeremiah Scharf, Christine Klein, Michael D Fox, R. Mark Richardson, Andreas Horn

## Abstract

Gene expression shapes the brain’s functional connectome, yet it is unclear whether genes linked to the same disorder converge on shared networks. We introduce gene network mapping–a framework combining spatial transcriptomics with normative functional connectivity to identify networks associated with gene expression. By generating *gene*-network maps, we captured distributed connectivity patterns for individual genes. Aggregating these across genes implicated in the same disorder yielded *disease*-network maps that captured the cumulative genetic impact on brain networks. We validated these maps by comparing them to lesion-derived networks and testing whether modulation of these networks predicted outcomes in deep brain stimulation (DBS) cohorts. This framework offers a novel tool to study the molecular architecture of brain disorders and supports the network-informed diagnostics and therapeutics in precision medicine.

## INTRODUCTION

The human brain is a complex organ that serves as the foundation of our thoughts, emotions, memories, and actions. Its intricate network of neural connections is fundamentally shaped by gene activity(*1*). Genes play a pivotal role in shaping the brain’s cortical and subcortical architecture(*2*), sculpting its networks(*3*), and orchestrating neural activity during brain development and throughout the human lifespan(*4–7*). Under physiological conditions genes promote growth, migration and differentiation of neurons, determine axonal trajectories, and regulate the assembly of synaptic connections including neurotransmitter systems(*2*). Recent studies have further expanded this concept by identifying gene regulatory networks that orchestrate brain-wide expression levels associated with behavioral responses(*8*). This accommodation enables neural plasticity, allowing the brain to rewire itself as needed to adapt to ever-changing external conditions.

Unfortunately, the same genetic mechanisms that form healthy brain networks may also lead to pathological circuit activity when disrupted(*9*). Genetic mutations, epigenetic modifications, and environmental factors can alter normal gene activity, leading to aberrant brain structure and function. Phenotypically, these changes can manifest as a wide array of neurological and psychiatric disorders, where abnormal gene expression and the subsequent dysfunction of brain circuits are causal in the onset and progression of these conditions. As a result, understanding the relationship between genes and brain networks carries profound implications to identify not only physiological but also pathological mechanisms. Notably, previous studies have successfully recapitulated resting-state fMRI (rs-fMRI) networks based on measures of correlated gene expressions(*10–12*), emphasizing crucial links between transcription patterns and neurodevelopment(*13*, *14*). Mapping entire *sets of genes* that are implicated in a given disease onto the functional network architecture of the human brain, however, has not been attempted previously. For instance, it has not yet been systematically examined whether the full set of genes linked to the occurrence of Parkinson’s Disease share common effects on distributed brain networks.

Spatial transcriptomics, a powerful high-throughput technique, has recently been introduced to examine local gene expression patterns in a spatially resolved manner(*1*, *15*). The advent of bulk-tissue microarray analysis, such as that used in the Allen Human Brain Atlas (AHBA)(*15*), has made it possible to profile over 20,000 genes across 3700 sampling locations in the human brain. This enables a more comprehensive analysis of local gene expression *patterns* across the entire brain. It provides detailed snapshots of gene expression within specific brain regions, and many studies have effectively used this approach to explore *regional differences* in pathological conditions(*16*, *17*). Recent years have seen the development of increasingly sophisticated approaches that integrate neuroimaging data with post-mortem transcriptomic datasets such as the AHBA. For instance, Seidlitz et al. introduced morphometric similarity networks and demonstrated their association with gene co-expression patterns, offering insights into brain development on a mesoscale(*18*). Patel et al. and Wagstyl et al. utilized vertex-wise modeling and advanced harmonization techniques to analyze transcriptomic influences in clinical populations and across cortical layers(*19*, *20*). Burt et al. used partial least squares regression to identify gene expression that aligned with macroscale functional and structural gradients(*21*). Forsyth et al. integrated biophysical connectivity modeling with transcriptional data to identify candidate genes driving cortical changes in 22q11.2 deletion syndrome(*22*).

Some of these studies have begun to explore the relationship between regional gene expression and macroscale features of brain organization, including structural and functional gradients, that reflect distributed network architectures(*18–23*). Moreover, some of these studies examined how multiple genes act in synchrony – using techniques such as partial least squares regression(*21*), canonical correlation analysis(*23*), or weighted gene co-expression network analysis(*18*) – to capture system-level transcriptomic signatures. Indeed, understanding how genes converge functionally may be critical to understand their collective impact on the connectome. However, comparably few studies have investigated whether sets of genes, each associated with a specific disorder, map to *shared* network structures(*19*, *22*). Such *networks*, rather than isolated brain *regions*, are fundamental to understanding brain function and dysfunction(*24*). Furthermore, it remains to be tested whether such gene-derived disease networks could inform targeted interventions such as brain stimulation.

The functional connectome has emerged as a promising tool to map causal insights onto polysynaptic brain networks(*25*). By pooling data from large cohorts of healthy participants, normative rs-fMRI provides a means to map functional connections between different brain regions in the average human brain. Leveraging the functional connectome, tools such as Lesion Network Mapping (LNM) (*26*, *27*) and Deep Brain Stimulation (DBS) network mapping(*28*, *29*) have proven effective in linking local disruption of neural activity to distributed brain networks. Lesion Network Mapping identifies shared connectivity patterns across brain lesions that each cause a specific clinical. Indeed, many studies have demonstrated that lesions causing the same symptom do not necessarily affect the same brain *area* but often connect to a shared, distributed brain *network*(*27*). DBS network mapping applies the same concept to stimulation volumes modeled from electrode reconstructions. Studies have demonstrated that distinct stimulation sites that improve the same symptom tend to fall into the same distributed brain network(*29*, *30*).

Building on these two frameworks, we propose that this approach could be applied to gene expression patterns to uncover networks associated with physiological or pathological brain states. Just as focal brain lesions or stimulation sites can have widespread effects beyond their anatomical location, genes may exert distributed influences across brain networks. By incorporating functional connectivity, we aimed to capture how gene expression shapes brain-wide activity and assess whether genes linked to specific symptoms or disorders converge onto common functional networks.

To test this approach, we first relate gene expression activity to the macroscale functional architecture of the human brain, generating *gene network maps*. We then aggregate multiple maps of genes implicated in a given disorder to create *disease-network maps*. These maps are first compared to networks informed by brain lesions that led to the same or different symptoms. Next, we assess whether clinical outcomes in patients who underwent deep brain stimulation are influenced by the extent to which the gene-derived networks has been modulated.

## Results

### Gene-network Maps Reveal Distinct Functional Connectivity Patterns Across the Brain

Our first aim was to generate and validate gene-network maps that could identify the distributed functional brain networks associated with *individual* gene expression patterns (Fig. 1A-B.). We achieved this by assigning both functional connectivity patterns and gene expression values to a brain parcellation consisting of 430 regions. Each of these parcels was used as a seed in the normative functional connectome, resulting in 430 distinct connectivity fingerprint maps, which represent functional connectivity between the parcel and each other brain voxel (Fig.1A.).

**Fig. 1.**
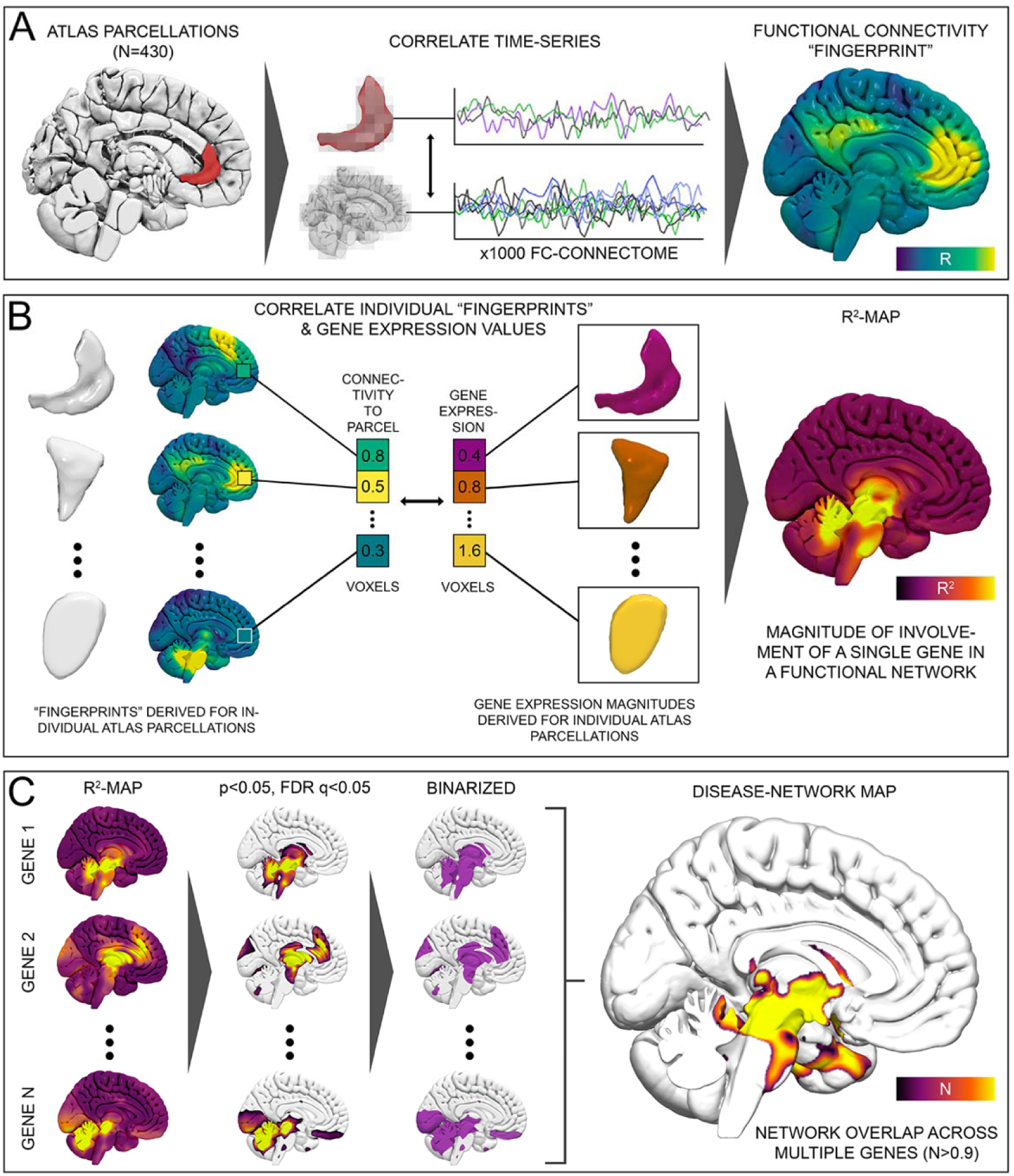
Methods used for the generation of gene- and disease-specific network maps. **(A)** Individual parcels of a whole brain labeling atlas (n=430 parcels) were used to seed blood-oxygen level dependent (BOLD) signal fluctuations from a normative functional connectome. The strength of correlation between seeded time series and every other voxel was calculated for each parcellation, producing a total of 430 functional connectivity “fingerprint” maps. **(B)** Expression magnitudes for a given gene were derived from the Allen Human Brain Atlas and assigned to each of the 430 brain parcels (three example parcellations of cingulate cortex, motor cortex, and pons are featured as well as their associated fingerprint maps and gene expression values). Linear models were then applied to determine the relationship between functional connectivity values and gene expression magnitudes on a voxel-by-voxel basis. This process yielded an R^2^-map, which we termed gene-network map, which captured the *magnitude* of the statistical relationship between gene expression and functional connectivity. **(C)** To identify common network connections across sets of genes associated with a specific system or disease, gene-network maps were thresholded, binarized, and merged. The ensuing disease-network map featured brain regions connected to a majority of relevant genes, revealing network hubs commonly implicated in gene networks. These maps provide insight into the shared functional connectivity patterns among genes within a particular disease context.

Next, we assigned expression magnitudes for an individual gene of interest derived from the AHBA to each of the 430 parcellations. The expression magnitudes in each parcel were then input into voxel-wise linear models using the connectivity values to the 430 brain parcels at a given voxel as independent variables and their gene expressions as dependent variable (Fig.1B.). The resulting gene-network maps represented R^2^ maps that quantified the relationships between gene expressions and functional connectivity to each voxel. Intuitively, these maps indicated which brain networks are most strongly associated with the overall expression patterns of a specific gene across the entire brain. A voxel with a high R2 value consequently indicated a brain region under pronounced influence of a given gene. The effect of using different brain atlas parcellations was minimal when this process was repeated (Fig. S1.).

Generating gene-network maps for different genes in the AHBA revealed a variety of functional connectivity patterns across the brain. Figure 2 highlights example maps for genes implicated in monogenetic disorders such as aromatic l-amino acid decarboxylase (AADC/DDC) deficiency, fragile X syndrome (FMR1), and dentatorubral-pallidoluysian atrophy (ATN1), respectively. Fitting with their pathophysiological relevance, the map associated with DDC predominantly highlighted a network that peaked in the substantia nigra, while FMR1 mapped to hippocampus, fornix, and amygdala(*31*),(*32*), and ATN1 mapped to the dentatorubral pathway and mesencephalon. We implemented two measures to validate the integrity of the approach. First, for each gene in the AHBA, we performed 10-fold cross-validation, recursively estimating gene expressions of left-out brain parcels based on gene-network maps generated from the remaining parcels. Cross-validation revealed the method’s capacity to produce reliable network models (average R: 0.49, range: −0.04 to 0.83). Second, we compared the distribution of R-values derived from cross-validation to a permutation-based null model. This analysis revealed statistical significance (P_perm_<0.001, two-sample t-test, two-sided tests), confirming that the associations observed in gene-network maps were not due to chance, but indeed represented meaningful relationships between the brain’s molecular architecture and function.

**Fig. 2.**
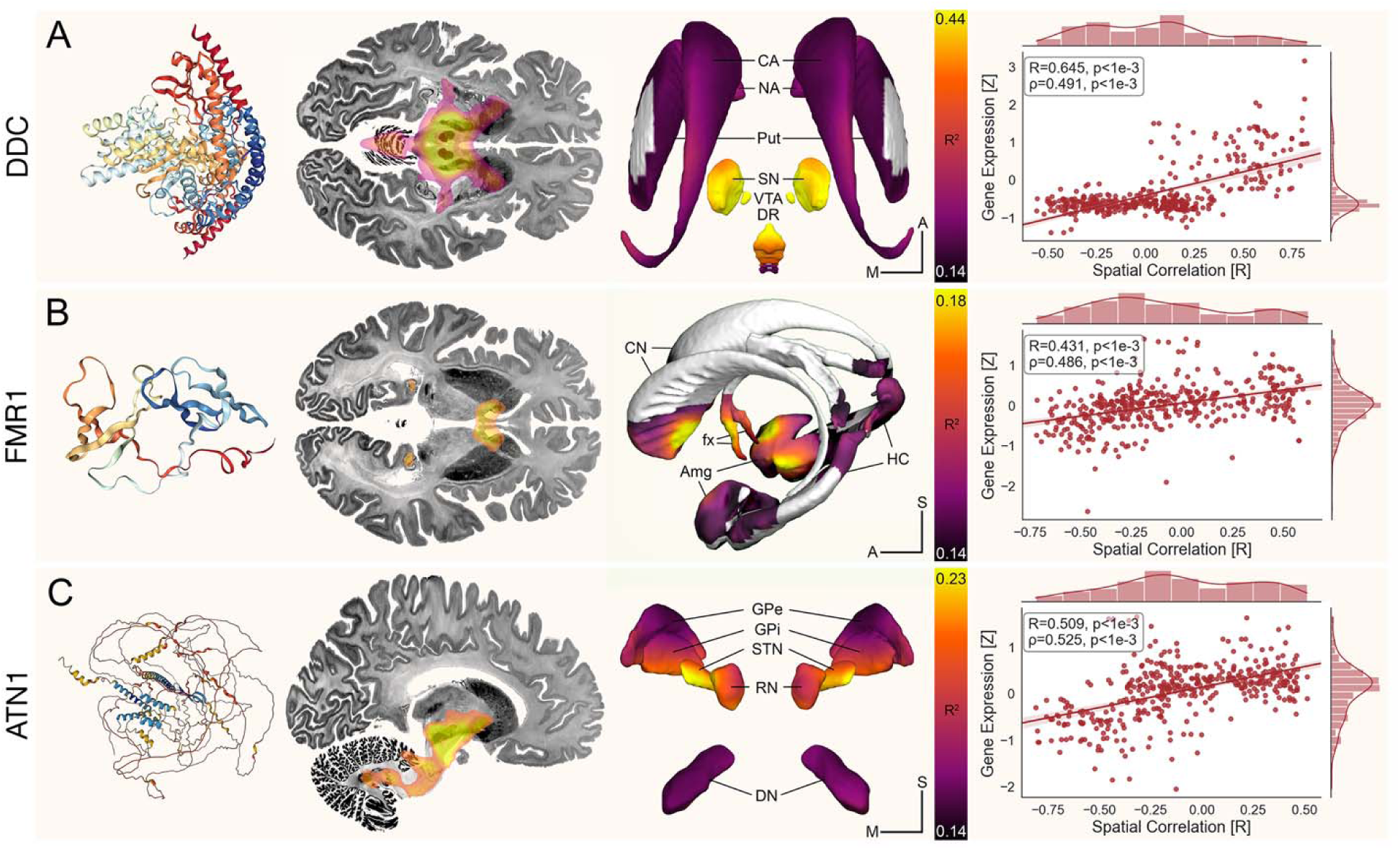
Validation of Gene-Network Maps. Example genes (rows) are shown with their corresponding protein structures as derived from the AlphaFold Protein Structure Database (https://alphafold.ebi.ac.uk). Heatmaps represent the association between functional connectivity (seeding from 430 parcels) and gene expression (expressed in each parcel). Peak intensities highlight the magnitude of the statistical relationship between gene expression and functional connectivity. The greatest involvement of a gene within a network is displayed at the cortical and subcortical level. Results are displayed against slices of the Big Brain Atlas(*33*) as well as subcortical structures extracted from the DISTAL Atlas(*34*) and CIT168 Atlas(*35*) for anatomical reference. Scatter plots feature the validation of gene-network maps using 10-fold cross-validation. Here, iteratively, gene-network maps were calculated based on a subset (N = 430 – 43, where 43 reflects the number of parcels in one of ten folds) of parcels and the resulting map was spatially correlated (x-axis) to functional connectivity fingerprints of all remaining parcels (N = 43, where 43 reflects the number of parcels in one of ten folds). These metrics were correlated with gene expressions in the remaining parcels (y-axis). Amg, Amygdala; CA, caudate nucleus; CN, caudate nucleus; DN, dentate nucleus; DR, dorsal raphe nuclei; fx, fornix; GPe, globus pallidus, pars externa; GPi, globus pallidus, pars interna; HC, hippocampus; NA, nucleus accumbens; Put, putamen; RN, red nucleus; SN, substantia nigra; STN, subthalamic nucleus; VTA, ventral tegmental area.

### Gene-Network Maps Capture Functional Connectivity Patterns Altered by Pharmacotherapy

On a single-gene level, cross-validation across parcels (above) demonstrated that gene network mapping can generate predictive models but left open the question of whether the approach captures genuine gene-network effects beyond statistical validation. To address this, we performed several analyses using pharmacological MRI (phMRI) and positron emission tomography (PET) data(*36*, *37*). Given that phMRI captures the functional consequences of neurotransmitter receptor engagement following pharmacological perturbation, we hypothesized that gene-network maps would be able to recapitulate connectivity patterns observed in the phMRI data. Conversely, we hypothesized that PET imaging, a local measure of neurotransmitter receptor and transporter density, would demonstrate less alignment with gene-network maps.

Confirming our hypotheses, results indicated that maps derived from genes implicated in neurotransmitter synthesis, regulation, and function reliably captured connectivity patterns induced by pharmacological agents (R=0.46, p<0.001 / ρ=0.46, p<0.001; Fig. S2.) and PET (R=0.35, p<0.001 / ρ=0.34, p<0.001; Fig. S3.). Additional control analyses that compare these relationships to local gene expressions and control for shared variance introduced by the functional connectome are detailed in the Supplementary Material. In summary, these findings supported the concept that the proposed method reliably captured network signatures, rather than localized brain regions (Supplementary Material).

### A syndrome-level View of Gene Expression-derived Functional Networks

The previous analysis demonstrated that functional networks derived from individual gene expressions can generate meaningful models. However, these analyses focused on single genes and did *not* address a pivotal question: how do *multiple* genes, associated with the same clinical syndrome or disorder, map to the topography of functional brain networks?

While some disorders are caused by mutations in a single gene, many traits and susceptibilities emerge from the combined effects of multiple genes on neural networks(*38*, *39*). By aggregating pathogenic variants of single genes that cause the same disease, we could identify common network substrates shared by those genes. Similarly, when applied to genes associated with a particular trait, this approach allowed us to map how their combined influence affected brain networks. In both cases, the method could help approximate the collective impact that segregated genes may exert on brain networks. Critically, this could theoretically work despite genes being spatially (expressed in different brain regions) and functionally (part of a distinct molecular pathway) segregated. Despite these differences, genes linked to shared symptoms may nonetheless converge on common networks – a hypothesis that remained to be tested.

To explore this, we identified common connectivity patterns across gene-network maps derived from *multiple* genes associated with a specific symptom or syndrome. Each gene-network map was thresholded using FDR correction (q<0.05) and binarized (Fig. 1C.). These binarized maps were then overlapped to create a group map, which we refer to as a *disease-network map*.

Although the conceptual framework of this approach appeared promising, the optimal method to validate the resulting maps was initially uncertain. Specifically, it was unclear which brain regions would emerge when generating a disease-network map from a set of predefined genes. To test the approach, we hence focused on movement disorders, specifically parkinsonism and dystonia, as their genetic underpinnings are well characterized(*40*, *41*) and their pathophysiological correlates well established(*42*), which allowed us to formulate testable hypotheses.

To identify high-probability genes associated with these disorders, we selected genes from the Movement Disorder Society (MDS) initiated Task Force for the Nomenclature of Genetic Movement Disorders, which offers comprehensive lists of genes associated with a given movement disorder(*43*, *44*) (table S1.). The resulting disease-network maps associated with parkinsonism and dystonia revealed distinct functional connectivity patterns across the brain (Fig. 3.). The parkinsonism map peaked in the mesencephalon, encompassing the substantia nigra and subthalamus. These connectivity patterns illustrated how genes with different spatial and functional expression profiles – peaking in various brain regions and involved in diverse and unrelated molecular pathways, like the ubiquitin-proteasome system, mitochondrial function, tyrosine metabolism, and axonal transport mechanisms – converged onto the nigrostriatal system, which is recognized as the primary anatomical substrate of the disorder(*45*). Over 90% of genes associated with parkinsonism further connected to a broader parkinsonian network that extended beyond the midbrain, including the basal ganglia, thalamus, deep cerebellar nuclei, and pons (Fig. 3.).

**Fig. 3.**
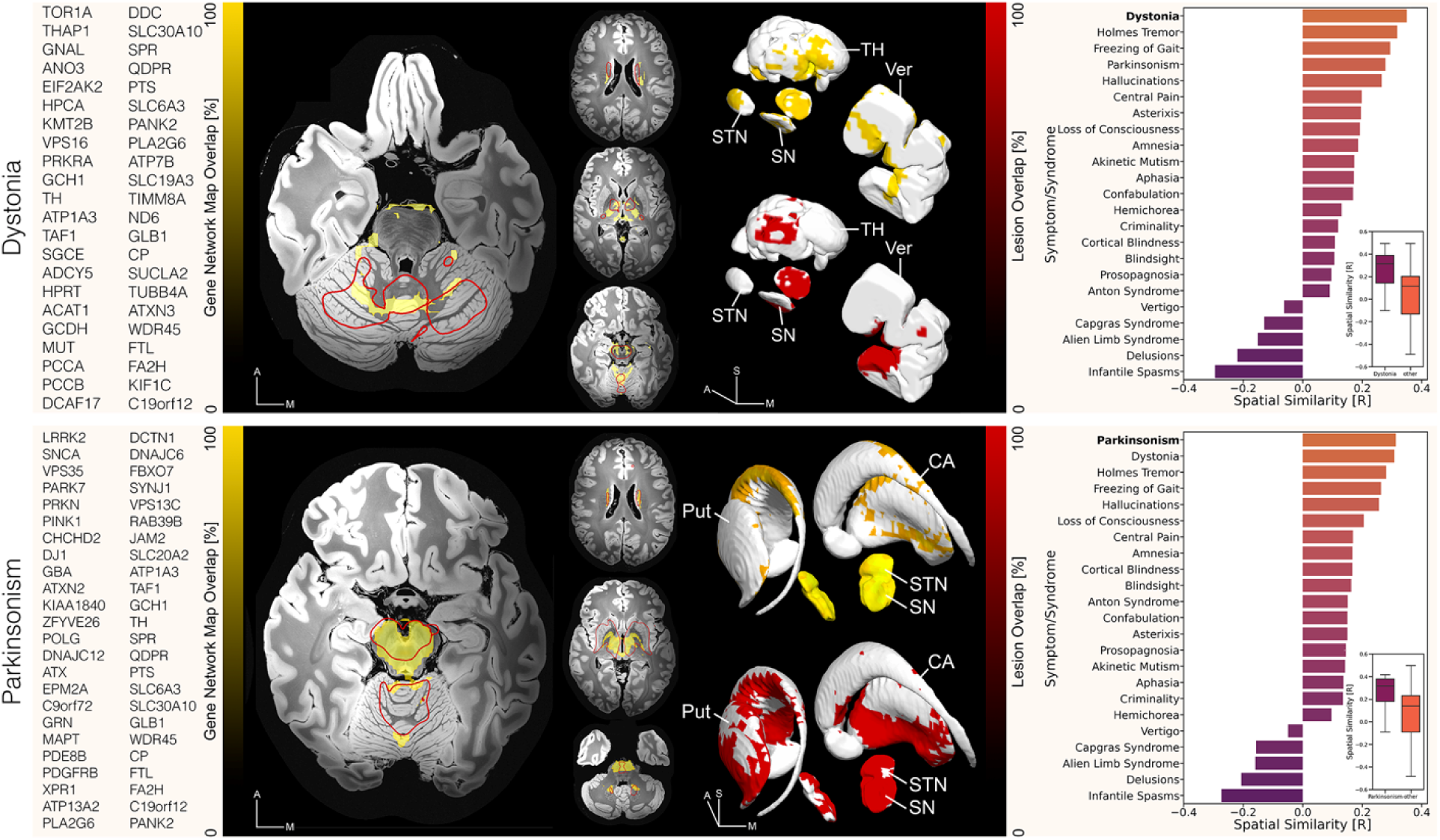
Spatial relationship between disease-network maps and Lesion Network Maps (LNMs) derived from brain lesions associated with dystonia and parkinsonism. Genes associated with each disease were derived from the Movement Disorder Society (MDS) initiated Task Force for the Nomenclature of Genetic Movement Disorders(*43*, *44*). Disease-network maps associated with dystonia (top row) and parkinsonism (bottom row) are superimposed on top of a 100-µm, 7T brain scan in MNI space(*51*) and are featured in yellow (n>0.9). The black-to-yellow color map identify common network substrates shared by individual genes, with yellow color indicating peak overlap. Lesion network maps associated with each disease are defined by red outlines. Overlapping peak regions across both disease and lesion-network maps emerge at their intersection. The black-to-red color map represents the degree of overlap between lesions causing the same disorder, i.e., dystonia (upper panel) or Parkinson’s disease (lower panel), with red color indicative of high lesion overlap. Additionally, both maps are featured in anatomical reference to subcortical atlas structures from the DISTAL atlas(*34*) and CIT168 atlas(*35*) to highlight subcortical peak regions. The right column features the spatial correlation of disease-network maps with connectivity profiles seeding from individual lesions associated with dystonia and parkinsonism and all other lesions from the Harvard lesion repository. Bar plots indicate spatial similarities stratified by symptom. Bars represent medians of similarity indices. Box plots feature the distribution of correlations of disease-network maps and disease-specific lesion profiles (purple) vs all other lesion profiles contained in the Harvard lesion repository (orange). CA, caudate nucleus; Put, putamen; SN, substantia nigra; STN, subthalamic nucleus, TH, thalamus; Ver, vermis.

A similar convergence was observed in the disease-network map generated for dystonia (Fig. 3.). Here, connectivity analysis identified the cerebellum, globus pallidus, midbrain and thalamus in over 90% of genes, all regions involved in treatment of dystonia(*46*, *47*). The largest cluster entailed the cerebellar vermis, which plays a significant role in movement coordination, posture, and balance and has also been implicated in the pathophysiology of dystonia(*48*).

### Lesional and Genetic Causes of Movement Disorders Converge on Common Networks

The two disease-network maps identified regions known to play a role in pathological contexts. However, since the data was derived from physiological gene expressions, it remained unclear whether our methodological choices could reliably identify pathological networks. Lesion Network Mapping, by contrast, represents an already well-validated approach that yields a high degree of causal insight into the functional connectivity landscape of the brain’s function(*49*, *50*). Brain lesions typically produce significant and observable behavioral, cognitive, or emotional changes that follow a clear temporal sequence after the lesion occurs. By mapping the effects of anatomically dispersed brain lesions onto a unified functional brain network, an unequivocal link between specific networks and particular symptoms or syndromes can hence be established, which cannot be explained by mere correlation or coincidence. To validate the utility of our mapping approach, we thus compared our genetic disease-network maps to network maps generated from brain lesions associated with the same disorders versus a total of 971 control lesions from the Harvard Lesion Repository(*50*) (Fig. 3., table S2.).

This analysis revealed greater similarities between lesion network maps and gene-derived disease-network maps corresponding to the same syndrome (i.e., parkinsonism and dystonia, respectively; Fig. 3.) compared to lesions related to other symptoms. Indeed, the disease-network map for parkinsonism demonstrated greater similarity with lesions causing parkinsonism than with lesions from 23 other syndromes in the database. The same was true for dystonia: lesions resulting in dystonia fell more strongly into the disease-network map for dystonia than lesions from 23 other syndromes.

### Disease-Network Maps Predict DBS-Induced Symptom Improvement in Dystonia and Parkinson’s Disease

While the observed similarity between gene networks and lesion sites associated with shared symptoms was encouraging, it remained unclear whether the identified disease-networks could inform treatment strategies, such as deep brain stimulation. We hypothesized that modulation of these networks through invasive stimulation would yield maximal symptom improvement for each respective disorder.

To test this hypothesis, we retrospectively analyzed a cohort of 118 DBS patients, with a total of 236 electrodes implanted to treat dystonia (n=39) and Parkinson’s disease (n=79) (table S3.)(*28*, *52–59*). We spatially correlated connectivity profiles of DBS stimulation volumes with genetic disease-network maps for each condition to determine how strongly the stimulated network aligned with the gene-derived target network. These analyses demonstrated that stimulating networks that closely aligned with the gene-derived disease-networks associated with optimal treatment response in both dystonia (R=0.56, p=6e-4/ ρ=0.44, p=0.008, Fig. 4.) and Parkinson’s disease (R=0.47, p=1e-5/ ρ=0.43, p=9e-5, Fig. 4.). Furthermore, genetic disease-network maps calculated for one condition associated poorly with DBS outcomes for the respective other condition (dystonia based on Parkinson’s disease genes: R=0.14, p=0.42/ ρ=0.18, p=0.32; Parkinson’s disease based on dystonia genes: R=0.28, p=0.01/ ρ=0.31, p=5e-3).

**Fig. 4.**
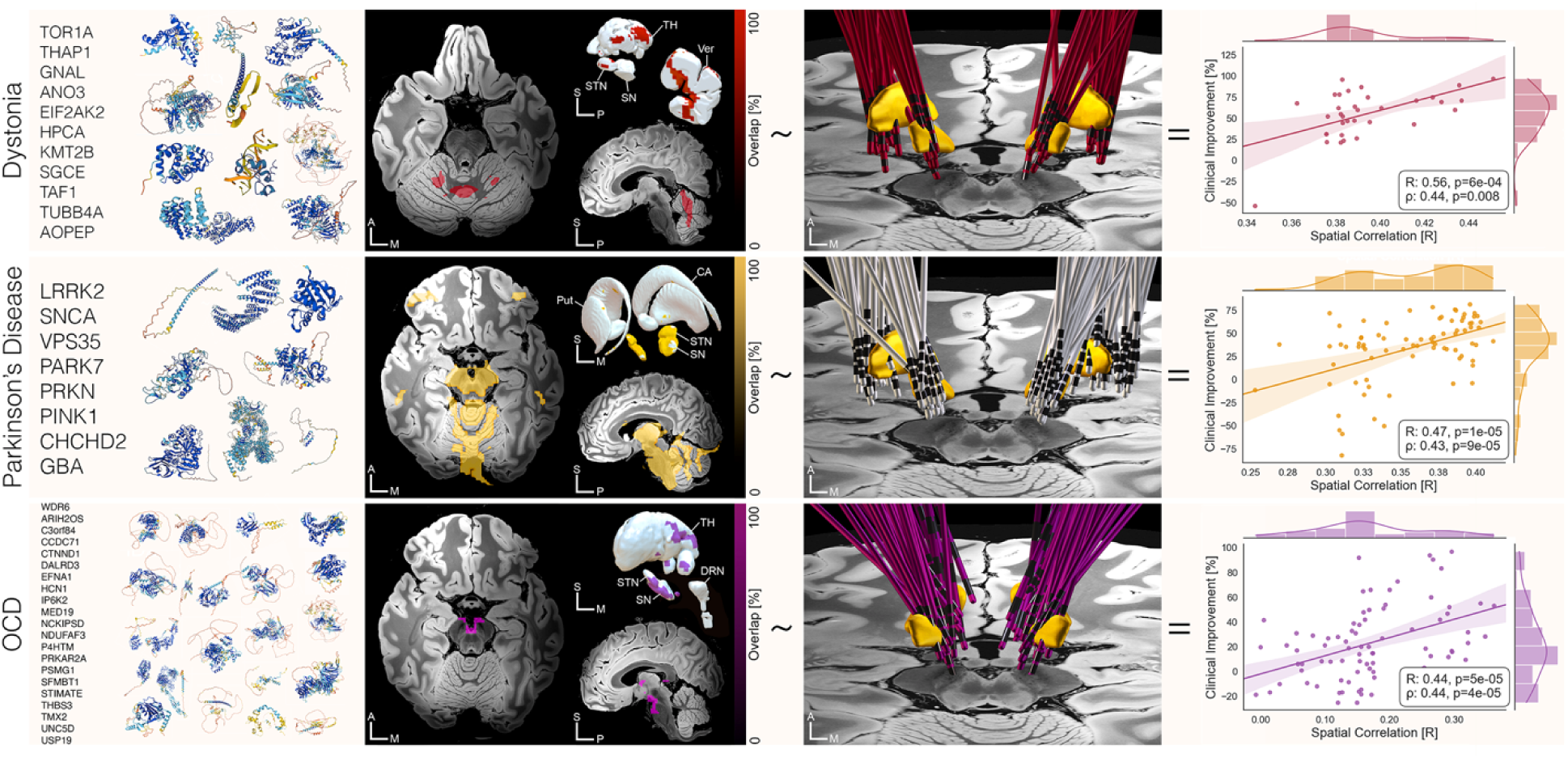
A genetic link between movement disorders, psychiatric diseases, and DBS outcomes. We generated disease-network maps for dystonia and Parkinson’s disease based on the spatial expression profile of genes that have been associated with symptom improvement following deep brain stimulation (DBS) surgery (top rows). To derive genes associated with obsessive-compulsive disorder (OCD), we relied on the largest case-control cohort analyzed to date (bottom row)(*60*). Maps were calculated by aggregating gene network maps of individual genes into disease-network maps. These maps, represented by the degree of overlap between individual gene-network maps, are superimposed onto a 100-µm, 7T brain scan in MNI space(*51*) (second column), with peak intensities featured in red (dystonia), yellow (Parkinson’s Disease), and violet (OCD), respectively. Maps are thresholded at n>90%. The third panel illustrates DBS electrode reconstructions across disorder-wise cohorts against relevant anatomical structures extracted from the DISTAL atlas(*34*) and the CIT168 atlas(*35*). Spatial correlation of disease-network maps with patient-specific connectivity maps seeding from stimulation volumes significantly predicted outcome across indications (right column). Optimal clinical outcome was closely tied to modulation of networks implicated in each disease’s pathophysiology, underscoring the potential of integrating genetic and neuroimaging data to guide DBS targeting and programming. CA, caudate nucleus; DRN, dorsal raphe nucleus; Put, putamen; SN, substantia nigra; STN, subthalamic nucleus, TH, thalamus; Ver, vermis.

Repeating the analyses using the broader, more inclusive set of genes associated with dystonia and parkinsonism (Fig. 3.), significant correlations persisted. However, the variance explained was notably reduced, particularly in parkinsonism (dystonia: R=0.59, p=3e-4/ ρ=0.45, p=7e-3; parkinsonism: R=0.21, p=0.022/ ρ=0.19, p=0.038). These results support our hypothesis that the narrower gene set associated with DBS-responsive conditions may yield a better representation of the diseased network that can be effectively modulated by DBS. Taken together, these findings underscore the potential of integrating genetic and neuroimaging data to guide DBS targeting and programming toward the correct circuits, which could potentially help to improve therapeutic outcomes. In a final analysis we sought to evaluate the applicability of our methodological approach to a psychiatric disorder. While it has been established that genetic factors play a role in the susceptibility to and development of Obsessive-Compulsive Disorder (OCD)(*61–63*), the disorder results from a complex interplay of genetic and environmental influences, and specific causal genes remain unclear. Indeed, genome-wide association studies (GWAS) of OCD published to date are significantly underpowered, resulting in less reliable gene identification in this patient population(*64–66*). Furthermore, GWAS can only identify genetic risk, no causal genes, although there may be overlap. We thus relied on previous work by Strom et al. to identify the most *probable* set of gene candidates increasing genetic risk in the context of OCD(*60*). In a substantially expanded case-control cohort (n=37.015 OCD cases vs n=948.616 controls), this work had identified 21 key genes significantly associated with OCD (table S1.). These genes produced a disease-network map with peak intensities in distinct limbic hubs of the subcortex including the ventral tegmental area (VTA), dorsomedial nucleus (DM) of the thalamus, ventral pallidum, and – to a lesser extent – the dorsal raphe nucleus (DRN; Fig. 4.). As in PD and dystonia, the degree of stimulation of this gene-based disease-network correlated with clinical improvements in a total of 160 DBS electrodes implanted in patients with therapy-refractory OCD (R=0.44, p=15e-5/ ρ=0.44, p=4e-5; Fig. 4., see table S3 for a detailed overview of included patient cohorts)(*55–59*).

## DISCUSSION

We introduce and apply a novel method termed gene network mapping, which integrates spatial transcriptomics with functional connectomics to identify *gene-* and *disease-level* networks that associate with gene expression. We demonstrated the applicability of this method to single genes (gene-network maps) and groups of genes associated with specific syndromes (disease-network maps). We subjected our framework to extensive validation analyses using a variety of approaches, including pharmacological MRI, PET imaging, Lesion Network Mapping, and DBS network mapping. Gene-network maps derived from neurotransmitter genes recapitulated network properties identified by both pharmacological MRI and PET. Aggregating sets of genes that formed a pathological system pointed to established brain regions known to play a pivotal role in the disease mechanism. Specifically, we showed that network maps generated from genes associated with parkinsonism and dystonia matched those from naturally occurring brain lesions causing the respective condition. Finally, our framework demonstrated a potential for clinical application, as disease-network maps could account for variance in symptom improvements in patient cohorts that underwent DBS for Parkinson’s disease, dystonia, and OCD. This finding underscores the translational promise of the approach, which may include gene-network-derived diagnostic modalities and therapeutic strategies.

The release of the Allen Human Brain Atlas (AHBA) represented a major breakthrough in human neuroscience of the present century(*15*). Not only did it mark a significant advancement in spatial transcriptomics, but also gave researchers the unprecedented opportunity to investigate the genetic underpinnings of the human brain. Preprocessing toolboxes, such as *abagen* further popularized the dataset as they provided robust and reproducible means for preprocessing microarray expression data derived from the AHBA(*67*, *15*, *50*, *68*). Consequently, studies leveraging these tools were able to identify links between dominant modes of gene expression and brain function, and recapitulated rs-fMRI networks based on measures of correlated gene expressions(*10*– *12*), emphasizing interplay between transcription patterns and neurodevelopment(*13*, *14*). Moreover, they established key associations with functional(*10*, *69*, *70*) and structural(*71*, *72*) connectivity phenotypes as well as neurotransmitter receptor densities(*73*). Further work associated molecular signatures with cognitive processes(*74*) and demonstrated links to pathological states, including neurodevelopmental(*75*, *76*), neurodegenerative(*77*), and psychiatric diseases(*78*, *79*).

Here, we mapped individual genes as well as sets of genes linked to brain disorders onto the functional network architecture of the human brain. At the disease-network level, our method aimed to approximate critical network hubs that linked to the majority of genes that associated with a given disorder. Using parkinsonism, dystonia, and – in an exploratory manner, OCD – as examples, we identified broader networks related to genes involved in these diseases. These connectivity patterns reflect the integration of genes with spatially and functionally distinct expression profiles and represent a convergence of genes with peak expressions in different brain regions and unrelated molecular pathways. Conceptually, the framework used for data analysis aligned with the Lesion Network mapping approach. This technique seeks to identify brain networks that commonly associate with brain lesions that cause the same symptom(*50*). Indeed, disease-network maps generated from parkinsonism and dystonia genes matched those derived from naturally occurring brain lesions causing the same symptoms (Fig. 3.). This alignment supports the hypothesis that networks disrupted by lesions may reflect those altered by pathogenic variants and not only validates the proposed gene-network approach, but also suggests a fundamental connection between network disruptions caused by *brain lesions* and *genetic mutations*. Moreover, the general finding aligns with recent work in dystonia(*80*), tics(*81*), and mania(*82*) which established similarities between networks identified through brain lesions and idiopathic causes of the disease. For example, *Corp et al.* showed that the cerebellar and somatosensory regions identified by LNM exhibited abnormal rs-fMRI connectivity in an independent dataset of idiopathic dystonia patients(*80*). Similarly, Lee *et al.* demonstrated that mania caused by brain lesions and mania as a symptom of bipolar disorder involves shared networks(*82*). Combined with the present findings, this suggest that lesion-induced, idiopathic, and genetic reasons that lead to the same symptom may often map to similarly distributed global brain networks.

While the impact of genes on brain pathology is well recognized, translating these insights into clinical applications remains a challenge. For example, although genes causally involved in Parkinson’s disease have been well established(*43*, *44*), selection criteria for DBS surgery are typically based on clinical features (e.g., motor fluctuations, absence of dementia or other psychiatric conditions), refractoriness to pharmacotherapy, and neuroimaging findings. Instead, while significant efforts are currently underway to improve the genetic characterization of DBS surgery candidates(*83–86*), genetic carrier status is currently not the focus in surgical decision-making for Parkinson’s Disease(*87*). Our results support further investigation into the potential of using genetic information in combination with current screening approaches to guide neuromodulation therapies. Namely, we compared network maps seeded from DBS electrodes in the treatment of Parkinson’s disease, dystonia, and OCD with disease-network maps based on genes associated with each respective disorder. For all three conditions, symptom improvement post-DBS significantly correlated with how closely the DBS-engaged network map resembled the genetic disease-network map. Building upon this, by incorporating patient-specific genetic carrier status, our approach could be refined to personalize treatment by emphasizing (up-weighing) networks associated with the identified mutations, potentially using metrics like polygenic risk scores to understand network effects at the individual level and guide screening and/or therapy(*88*, *89*). Owing to a lack of genetic information in the analyzed DBS cohorts, such analyses were not possible in the present study.

The example of DBS demonstrates that gene-network mapping as the integration of spatial transcriptomics and functional connectivity may constitute a promising framework for clinical translation that could be applied to several other concepts. First, the method may identify candidate genes that could contribute to disease etiology, providing new avenues for genetic screening and gene therapy. For example, connectivity patterns of putative causative genes and their associated networks may be referenced against existing disease-network maps to accelerate identification of candidate genes. Second, the framework may allow mechanistic insights into the pathophysiology of brain disorders. Al-Fatly et al. previously demonstrated this potential in the case of Oculogyric Crises where, the pathological network derived from brain lesions causing this disorder revealed spatial overlaps with genes coding for dopamine receptors type 2 and 3 (DRD2/DRD3), providing insight into the relationship between receptor distributions and lesion-induced network disruptions(*90*). Furthermore, while not explored here, gene-network mapping may offer a promising platform for high-throughput screening in drug discovery and drug repurposing. By modeling the effects of potential compounds on gene networks, we may be able to predict their impact on disease-related neural circuits and refine drug candidates before clinical trials. This strategy not only has the potential to accelerate and reduce cost in the process of drug discovery but may also increase the likelihood of success in clinical applications by ensuring that drugs target relevant brain networks. Finally, this approach could be instrumental in overcoming the challenges associated with polygenic diseases, where the cumulative effect of multiple genes complicates the identification of therapeutic targets. By focusing on network-level disruptions rather than individual genes, gene-network mapping may allow us to capture the broader landscape of disease pathology, leading to more effective and comprehensive treatment strategies.

Despite generally promising results, this study has several limitations that should be acknowledged. First, it is important to note the general limitations associated with the use of the Allen Human Brain Atlas. While being the most spatially comprehensive data repository currently available for gene expression analysis in the human brain the number of samples is low and, like other transcriptomic datasets, it is derived from postmortem brains, which may not fully reflect gene expression in living tissue(*91*). Preprocessing of microarray expression data has historically posed challenges due to variable preprocessing pipelines. However, by employing abagen(*68*), a preprocessing toolbox for the AHBA dataset, we were able to mitigate many of the problems associated with the original dataset ensuring a standardized workflow consistent with current best practices. Second, our analysis inferred pathological gene network activation from physiological expression levels and functional connectivity data. Given the variety of downstream effects that changes in gene expression may have on metabolic pathways and functional networks, we avoided assumptions about the directionality of the observed effects during system network map generation, focusing instead on the magnitude of the relationship between genes and functional connectivity. This approach seeks to provide a balanced perspective, particularly regarding the cumulative effects of pathological genes on a network, though it may not capture the full complexity of gene-network dynamics in disease states. Further research is required to validate this analytical choice. Third, it is important to note that genotyping was not performed in the DBS patients investigated here, and the pathology in the majority of patients was idiopathic or associated with unknown genetic or epigenetic modifications. However, the overlap between patient-specific DBS-outcome maps and disease-network maps, explained variance in outcomes across a heterogeneous cohort. This provides further evidence that our disease-network maps may capture fundamental aspects of disease pathology irrespective of the underlying pathogenic cause, as was also demonstrated with Lesion Network Mapping. Finally, our modeling relied on genes causally associated with each respective disease. The exception was OCD, where we relied on candidate genes identified by the largest genome-wide association study of OCD to date(*92*). While the approach still led to significant predictions of DBS outcomes in patients with OCD, it remains to be seen how well it performs when incorporating genes of even lower confidence. It is theoretically possible that aggregating low-confidence genes could result in a network of high confidence, but alternatively, the ability to explain variance may be reduced if network effects are obscured by noise. Future research is warranted to explore ways of improving the signal-to-noise ratio in such cases, potentially by personalizing the modeling approach using polygenetic risk scores.

## CONCLUSIONS

Our study introduces a novel method for integrating the human transcriptome with the human connectome, enabling us to comprehensively map functional networks associated with individual genes and, critically, *sets* of genes. By validating our approach against established methods such as pharmacological MRI, PET, Lesion Network Mapping and DBS network mapping, we demonstrated robustness and potential for translation. This work may open new avenues for personalized medicine, with the promise of tailoring therapeutic interventions, including gene discovery, drug discovery, pharmacotherapy, and neuromodulation based on an individual’s genetic and network profile. As we advance, the integration of multiscale molecular and phenotyping approaches will be crucial for refining these strategies and improving patient outcomes.

## Data Availability

All data produced in the present study are available upon reasonable request to the authors

## ACKNOWLEDGMENTS

The authors are grateful to Matthew State for helpful general advice and feedback on the present work. They also thank the patients and their families for their participation in the clinical trials that enabled collection of the invaluable data analyzed here.

## FUNDING

Einstein Center for Neurosciences Berlin scholarship (BH)

Benign Essential Blepharospasm Research Foundation (JLO, PAS)

Institutional funds from Scuola Superiore Sant’Anna (MV)

National Institute for Health Research (NIHR)

University College London Hospitals Biomedical Research Centre (HA)

German Research Foundation (DFG) grant TRR 295 – Project-ID 424778381 (AAK, AH, NL)

German Research Foundation (DFG) grant Project-ID 347325977 (AAK)

Medical Research Council UK grant MR/J012009/1 (EMJ, LZ)

National Institutes of Health grant R01MH113929 (MDF)

National Institutes of Health grant R21MH126271 (MDF)

National Institutes of Health grant R56AG069086 (MDF)

National Institutes of Health grant R21NS123813 (MDF)

National Institutes of Health grant R01NS127892 (MDF, AH)

Kaye Family Research Endowment (MDF)

Ellison/Baszucki Family Foundation (MDF)

Manley Family (MDF)

German Aerospace Center (DLR)

DynaSti grant within the EU Joint Programme – Neurodegenerative Disease Research (AH)

National Institutes of Health grant R01 13478451 (AH)

National Institutes of Health grant 1R01NS127892-01 (AH)

National Institutes of Health grant 2R01MH113929 (AH)

New Venture Fund FFOR Seed Grant (AH)

BRAIN Initiative CONNECTS comprehensive center award UM1-NS132358 (AH)

## AUTHOR CONTRIBUTIONS

Conceptualization: CN, AH

Methodology: CN, BAF, NL, KB, MV, AB, NDS, ET, FS, BB, AH Software: BAF, NL, KB, MV, AB, NDS, ET, FS, BB, AH

Investigation: CG, JS, CK, MDF, RMR

Resources: PM, HA, NA, DC, SR, PB, JLO, PAS, VJJO, RMAB, JAB, HT, SAS, WKG, VVV, MF, DDD, LZ, EJ, DC, JJ, TP, KF, AAK, BAF, BH, NL, GMM, NR, FS, LH, SM, JP, BAF, NL, KB, MV, AB, NDS, ET, FS, BB, CG, JS, CK, MDF, RMR, AH

Data curation: CN, PM, HA, NA, DC, SR, PB, JLO, PAS, VJJO, RMAB, JAB, HT, SAS, WKG, VVV, MF, DDD, LZ, EJ, DC, JJ, TP, KF, AAK, BAF, BH, NL, GMM, NR, FS, LH, SM, JP, BAF, NL, KB, MV, AB, NDS, ET, FS, BB, CG, JS, CK, MDF, RMR, AH

Formal analysis: CN, BAF, BH, NL, GMM, NR, FS, LH, SM, JP, AH

Visualization: CN, AH

Writing – original draft: CN, AH

Writing – review & editing: PM, HA, NA, DC, SR, PB, JLO, PAS, VJJO, RMAB, JAB, HT, SAS, WKG, VVV, MF, DDD, LZ, EJ, DC, JJ, TP, KF, AAK, BAF, BH, NL, GMM, NR, FS, LH, SM, JP, BAF, NL, KB, MV, AB, NDS, ET, FS, BB, CG, JS, CK, MDF, RMR, AH

Supervision: AH

Project administration: AH Funding acquisition: AH

## DATA AND MATERIALS AVAILABILITY

The Allen Human Brain Atlas transcriptomic database is available at https://human.brain-map.org. The libraries used for preprocessing of the transcriptomic data can be freely accessed via GitHub (https://github.com/rmarkello/abagen). The PET raw data has been made publicly available and can be openly downloaded (https://github.com/netneurolab/hansen_receptors). Preprocessed pharmacological MRI scans are publicly available and can be downloaded as auxilliary dataset files (https://www.science.org/doi/10.1126/sciadv.adf8332). Detailed patient-wise demographic and clinical information is available in Supplementary Tables 2 and 3 in anonymized form. Patient imaging data cannot be publicly shared as this would compromise patient privacy according to current data protection regulations. They are, however, available from the principal investigators of the collecting sites upon reasonable request within the framework of a data-sharing agreement. Inquiries for further information and data-sharing requests should be directed to the corresponding authors of this manuscript (C.N., or A.H.) who commit to replying to any request within a timeframe of 30 days. Although a processed version of the 1000 GSP healthy subjects Connectomecan be requested from the corresponding authors, source data are freely accessible via the repository of the Harvard Dataverse (https://doi.org/10.7910/DVN/KKTJQC). The DISTAL atlas, version 1.1, and the CIT168 atlas, version 1.1, are openly available via the Lead-DBS knowledge base (https://www.lead-dbs.org/helpsupport/knowledge-base/atlasesresources/atlases-2/) and come pre-installed with the Lead-DBS software. The code used in the analyses presented in this work is openly available within the Lead-DBS environment (https://github.com/leaddbs/leaddbs).

## SUPPLEMENTARY MATERIALS

Materials and Methods

Supplementary Text

Figs. S1 to S3

Tables S1 to S3

References (1–115).

