## Supplementary Material for "Translating the Transcriptome: A Connectomics Approach for Gene-Network Mapping and Clinical Application"

(Neudorfer et al., 2025)

SUPPLEMENTARY MATERIAL

**MATERIAL AND METHODS**

**Gene selection**

To generate disease-specific network maps, we identified genes known to be causally involved in generating the disease phenotype. Genes associated with neurotransmitter systems were derived from the literature as described in Supplementary Table 1. Single-gene disorders implicated in distinct movement disorder phenotypes were derived from the Movement Disorder Society (MDS) initiated Task Force for the Nomenclature of Genetic Movement Disorders(*43*, *44*). For analysis, we focused on two syndromes, namely parkinsonism and dystonia, as their underlying causative pathogenic variants are well-established in the literature and have been comprehensively summarized by MDS. A detailed overview of gene selection and validation strategies can be found in Marras et al.(*43*) and Lange et al.(*44*). A complete list of genes employed in the present study can be obtained from Supplementary Table 1. For the correlation of disease-network maps with DBS network maps we used a subset of genes that was demonstrated to yield symptom improvement following DBS (Supplementary Table 1). Unlike single-gene disorders with clear causal links, the genetic underpinnings of OCD are complex and less certain featuring a highly polygenic architecture and correlations to other psychiatric phenotypes(*93*). Therefore, suitable gene candidates were identified by consulting with experts in the field and relying on previous work by Strom et al. to identify the most probable set of genes in OCD (Supplementary Table 1)(*60*).

**Preprocessing of the AHBA microarray expression data**

Regional microarray expression data were obtained from six post-mortem brains (1 female, ages 24-57 years) provided by the Allen Human Brain Atlas (AHBA, https://human.brain-map.org)(*15*). Using a 430-region whole-brain volumetric atlas in MNI space with approximately equal surface areas(*94*, *95*) we processed regional microarray expression data with the abagen toolbox (version 0.1.3; <https://github.com/rmarkello/abagen>)(*68*). First, microarray probes were reannotated using data provided by Arnatkevic̆iūtė et al.(*96*); probes not matched to a valid Entrez ID were discarded. Next, probes were filtered based on their expression magnitude relative to background noise, discarding probes with intensity less than the background(*97*). When multiple probes indexed the expression of the same gene, we selected and used the probe with the most consistent pattern of regional variation across donors determined by their differential stability(*1*), using the following formula:

$$\Delta_{S}\left( p \right)=\frac{1}{\binom{N}{2}} \sum_{i=1}^{N-1} \sum_{j=i+1}^{N} \rho\left[ B_{i}\left( p \right),B_{j}(p) \right]$$

where $\rho$ is Spearman's rank correlation of the expression of a single probe, p, across regions in two donors $B_{i}$and $B_{j}$, and N is the total number of donors. Here, regions correspond to the structural designations provided in the ontology from the AHBA.

The MNI coordinates of tissue samples were updated following coregistration using SPM12 (<https://www.fil.ion.ucl.ac.uk/spm/software/spm12/>) and nonlinear transformation into ICBM 2009b Nonlinear Asymmetric (MNI) space using the ANTs Symmetric Normalization (SyN) algorithm with the ‘effective: low variance + subcortical refinement’ preset as implemented in Lead-DBS v3.0(*98*, *99*). To increase spatial coverage, tissue samples were mirrored bilaterally across the left and right hemispheres(*71*). Samples were assigned to brain regions in the provided atlas if their MNI coordinates were within 2 mm of a given parcel. To reduce the potential for misassignment, sample-to-region matching was constrained by hemisphere and gross structural divisions (i.e., cortex, subcortex/brainstem, and cerebellum), such that – for example – a sample in the left cortex could only be assigned to an atlas parcel in the left cortex(*96*). All tissue samples not assigned to a brain region in the provided atlas were discarded. Inter-subject variation was addressed by mean- and variance-normalizing (i.e., z-scoring) tissue sample expression values across genes independently for each donor. Gene expression values were then normalized across tissue samples using an identical procedure. Samples assigned to the same brain region were averaged separately for each donor and then across donors, yielding a regional sample x gene expression matrix.

**Generation of gene-specific network maps**

To integrate gene expression values with functional networks, we adopted a methodological approach analogous to DBS network mapping as introduced by Horn et al.(*28*) Measures of functional connectivity were derived from high-resolution resting-state functional MRI (rs-fMRI) data sourced from the Brain Genomics Superstruct Project(*100*). Each of the 430 atlas parcellations was binarized and used to seed average blood-oxygen level dependent (BOLD) signal fluctuations from a total of 1,000 healthy participants. For each of the 1,000 normative brains, we then calculated the strength of the correlation between the seeded time series and every other brain-wide voxel outside the parcellation. These correlations were averaged across the normative sample and Fisher z-transformed to derive a normalized connectivity fingerprint for each parcellation. The subsequent steps were applied for each gene individually. Using linear models, we then modeled the relationship between fingerprint maps and gene expression values derived from each parcellation (n=430) in a voxel-wise manner. The resulting map indicates the strength of the relationship between gene expression levels and connectivity for each gene.

At the cellular level, mutations may have various downstream effects, including up- or downregulation of cellular processes such as protein synthesis, metabolic pathways, cell signaling, and cell cycle regulation(*101*). In a pathological context, a gene’s function and role within a biological pathway may be drastically altered. Mutation may cause either a gain of function, leading to new or abnormal gene activity that is harmful or improperly regulated, or a loss of function, resulting in the disruption of protective gene activity. At the network level, these changes may either enhance or attenuate functional connectivity, depending on the functions of the specific gene in question(*102*). Given the complexity of these gene-cell and gene-network interactions in the context of fMRI, and considering that the transcriptomic data was derived from physiological gene expression, we avoided assumptions about the *directionality* of the observed effect in these pathological cases. Instead, we captured the *magnitude* of the statistical relationship between gene expression and functional connectivity by generating R^2^-maps. In other words, R^2^ maps will simply indicate an association between gene activity and functional connectivity without specifying the directionality of the association.

**Generation of disease-specific network maps**

A derivative of the lesion network mapping approach was adopted to integrate multiple gene-specific network maps, thereby identifying brain regions consistently associated with individual symptoms across different genes(*50*). Gene-specific network maps were first thresholded and binarized using voxel-wise FDR correction (q<0.05). The resulting maps were then merged (i.e., summed and divided by the number of gene-specific network maps) to identify the common network connections across all investigated genes. From here onwards, we refer to these network overlap maps as *disease-network maps*. The value of each voxel in this map represents the proportion of gene-specific networks functionally connected to that voxel. Final maps were further thresholded to emphasize clusters of voxels connected to a majority of genes (n>90%) and discard brain regions that may have emerged through the contribution of individual outlier genes, in line with previously established methods(*103*).

**Validation of gene and disease-specific network maps**

We conducted several control analyses to assess the validity of our results and ensure that the observed findings were not specific to the initial sample and would generalize across different populations. The robustness of gene-specific network maps was evaluated using a 10-fold cross-validation approach. This method was used to predict the gene expression levels in brain regions excluded from the initial model construction to avoid the bias of circularity. Specifically, a gene-network map was generated from a subset of brain parcels during each of the ten folds of the analysis and spatially correlated with the functional connectivity patterns of the remaining parcels. After deriving estimates for each parcel across folds, spatial similarity values were correlated with gene expression values across all brain parcels to assess model validity. The correlation strengths obtained from this cross-validation for all genes was then compared against a control distribution using a two-sample t-test. Additional control analyses included i) generation of gene-network maps using a different set of atlas parcellations (defined by the Allen Human Brain Atlas(*95*) and the Mindboggle 101 Atlas(*104*), respectively), and ii) validations (see below) using independent datasets, ensuring that the observed results were not specific to the initial sample and would generalize across different populations.

**Validation**

**Pharmacological MRI**

To evaluate our methodological framework and compare the performance of generated gene-network maps to local expression patterns, we employed a publicly available dataset of pharmacological MRI (phMRI) data(*37*). This dataset comprises fMRI data from volunteers under the influence of 10 distinct pharmacological agents. These included psychedelics, such as ayahuasca, lysergic acid diethylamide (LSD), 3,4-methylenedioxymethamphetamine (MDMA), N,N-Dimethyltryptamine (DMT), psilocybin; cognitive enhancers, including methylphenidate and modafinil; and the anesthetics propofol, sevoflurane, and ketamine (administered in both, subanesthetic and anesthetic doses). In the latter, two contrasts were considered: drug vs. pre-induction baseline and drug vs. postanesthetic recovery. Data were acquired from a total of 224 participants across 382 fMRI sessions. For a detailed overview of the data acquisition, preprocessing, and validation processes, refer to(*37*). In brief, for each drug, the pattern of pharmacologically induced functional change was determined by comparing the within-subject differences in task-free baseline imaging to scans taken under the influence of each drug. The average change in regional functional connectivity across-subjects was then used to generate the final phMRI maps. These maps were parcellated into 100 functionally defined regions according to the Schaefer atlas(*37*, *105*).

**PET**

In addition to phMRI, we evaluated gene-network maps derived from neurotransmitter systems using a previously published reference database of PET tracer images, as curated by(*36*). The preprocessing of these data was conducted in line with the methodology described by(*37*) Receptor and transporter maps were compiled accounting for redundancy of scans associated with the same receptor/transporter and diversity of tracers associated with the same receptor/transporter. PET scans with more than one mean image of the same tracer (i.e., 5-HT1B, 5HTT, D2, MU, mGluR5, NATm, and VAChT) were averaged, weighing each image by the number of participants in each cohort. In cases where different tracers were used for the same receptor/transporter, our primary analysis utilized the tracer image with the largest patient cohort. Employing this approach, previous studies demonstrated consistent results across tracer subtypes(*73*).

**Comparison of Local Maps and Gene-Network Maps**

We generated covariance matrices based on various data types, including phMRI, local gene expression patterns, gene-network maps, and PET density measurements. For analysis, we used the full phMRI and PET datasets as introduced by(*36*, *37*); for local gene expression and gene-network maps we used gene transcripts coding for neurotransmitter systems as described by Hawrylycz et al.(*15*) (Supplementary Table 1). For each data type, we first parcellated the spatial patterns in the respective maps according to the Schaefer atlas(*105*). Then, within each data type, we correlated the concatenated parcel-wise distributions in a pairwise manner across all brain regions (see also Supplementary Material). This resulted in a set of covariance matrices reflecting the relationships and connectivity of the underlying maps within each specific data type. Pharmacological co-susceptibility was then correlated with the remaining covariance matrices in a pair-wise manner to determine the association of pharmacological co-susceptibility with local gene co-expression, gene-network co-activation, and PET co-localization (for more details refer to Supplementary Material).

**Lesions**

We determined whether the disease-specific network maps generated from transcriptomic data were sensitive to LNMs associated with the same symptom, but also specific to other naturally occurring brain lesions, not associated with the same symptom. To this end, connectivity T-maps derived from brain lesions causing parkinsonism (n=29) and dystonia (n=25) were compared to a total of 861 control lesions from the Harvard Lesion Repository(*50*). This repository contains lesions associated with various neurological and psychiatric symptoms including (numbers indicate lesion counts in each specific category): Akinetic Mutism, 28; Alien Limb, 53; Anton Syndrome, 24; Aphasia, 12; Asterixis, 30; Blindsight, 68; Capgras Syndrome, 17; Central Pain, 23; Confabulations, 25; Cortical Blindness, 70; Criminality, 17; Delusions, 15; Amnesia, 53; Freezing of Gait, 14; Hallucinations, 89; Hemichorea, 29; Holmes Tremor, 14; Infantile Spasms, 123; Loss of Consciousness, 28; Mania, 56; Prosopagnosia, 44; Vertigo, 23 (Supplementary Table 2).

Sensitivity and specificity were calculated by deriving the distribution of spatial correlations between the disease-specific network map and LNMs associated with the same symptom. The resulting distribution of R-values was then compared to a second distribution derived from spatial correlation of the same disease-specific network map with all other LNMs not associated with the symptom under investigation. Distributions were compared using a two-sample t-test. In addition, a stratification based on symptom type was performed.

**Deep Brain Stimulation**

To determine the relevance and potential clinical utility of the disease-specific network maps, we analyzed a total of 176 DBS patients across ten international DBS centers (Amsterdam, Berlin, Boston, Brisbane, Cologne, Grenoble, London, Madrid, Melbourne, and San Francisco) who underwent lead implantation for the treatment of dystonia (2 centers, n=39 patients), Parkinson’s Disease (2 centers, n=79 patients), and Obsessive-Compulsive Disorder (6 centers, n=58 patients)(*28*, *52*–*59*). All patients had been well characterized in prior retrospective trials (for patient demographics, clinical characteristics, and citations of respective publications refer to Supplementary Table 3). Lead implantation was performed across various targets including globus pallidus, pars interna (GPi) (dystonia, n=12; PD, n=51), subthalamic nucleus (STN) (dystonia, n=27; OCD, n=6; PD, n=28), ventral capsule/ventral striatum (VC/VS) (n=34), nucleus accumbens (n=15), and bed nucleus of the stria terminalis (n=9). In all patients, accurate lead placement was confirmed using either postoperative computed tomography (CT) or MRI. DBS leads were reconstructed using Lead-DBS v3.0(*98*). To this end, multimodal imaging of each patient was first co-registered using SPM12 and then nonlinearly transformed into MNI space using the ‘effective: low variance + subcortical refinement’ preset for ANTs. Localization of DBS leads was performed on postoperative CT or MRI scans following automatic (DYT and PD) and manual (OCD) pre-localization. Subsequently, stimulation volumes associated with patient-specific stimulation settings were modeled in native space using an adaptation of the SimBio/FieldTrip pipeline (<https://www.mrt.uni-jena.de/simbio/index.php/>; <http://fieldtriptoolbox.org>)(*106*) as implemented in Lead-DBS(*98*). Using a finite element modeling (FEM) approach, a volume conductor model was generated based on a tetrahedral four-compartment mesh, comprising gray matter, white matter, metal, and insulating electrode parts. The calculated stimulation volumes were then transformed into MNI space using the transforms generated during normalization and used to seed average blood-oxygen level dependent (BOLD) signal fluctuations from a normative rs-fMRI dataset of 1000 healthy participants as described earlier.

To determine whether networks associated with optimal outcome in the context of DBS show an association with transcriptome-derived disease-specific network maps, we generated disease-specific network maps from genes that had been associated with symptom improvement in dystonia, Parkinson’s Disease, and OCD in the DBS literature (Supplementary Table 1). We then calculated the spatial correlations between the disease-specific network maps and patient-specific fingerprint maps associated with the same symptom. Correlation between the derived distribution of R-values and clinical outcome of patients was then tested using Pearson and Spearman correlations. A p-value <0.05 was considered statistically significant.

**Statistical Analysis**

Statistical analysis was performed using MATLAB version 2023b (Mathworks) and Python 3.10.10. In this manuscript, Pearson correlation coefficients are reported as R, and Spearman correlation coefficients are denoted by ρ.

**Table S1**: List of resources used to investigate neurotransmitter systems and generate system- and disease-specific maps.

|  | phMRI | Neurotransmitter co-expression/ co-activation | PET | PET-matched genes | Dopamine System | Parkinsonism | Dystonia | Parkinson’s Disease (DBS) | Dystonia (DBS) | OCD (DBS) |
| --- | --- | --- | --- | --- | --- | --- | --- | --- | --- | --- |
| Reference | (*37*) | (*15*) | (*36*, *37*) |  | (*15*) | (*43*, *44*) | (*43*, *44*) | (*107*–*109*) | (*110*–*114*) | (*115*) |
|  | Ayahuasca  LSD  MDMA  DMT  Psilocybin  Ketamine  Methylphenidate  Modafinil  Propofol  Sevoflurane | TH; DDC PAH QDPR TYR DCT; SLC18A1; SLC18A2 CDNF GCH1 SPR TYRP1 CYP2D6 DAT; COMT PCBD1 PTS CALY PPP1R1B; SLC6A2; SLC6A3; DRD1; DRD2; DRD3; DRD4; DRD5; CHAT; SLC18A3; ACHE; BCHE; SLC5A7; SLC44A4; SLC44A1; SLC44A2; SLC44A3; SLC44A5; SLC22A2; SLC22A1; CHRM1; CHRM2; CHRM3; CHRM4; CHRM5; CHRNA1; CHRNA2; CHRNA3; CHRNA4; CHRNA5; CHRNA6; CHRNA7; CHRNA9; CHRNA10; CHRNB1; CHRNB2; CHRNB3; CHRNB4; CHRND; CHRNE; CHRNG; PNMT; GAD1; GAD2; SLC32A1; SLC6A1; SLC6A11; SLC6A13; ABAT; GABBR1; GABBR2; GABRA1; GABRA2; GABRA3; GABRA4; GABRA5; GABRA6; GABRB1; GABRB2; GABRB3; GABRD; GABRE; GABRG1; GABRG2; GABRG3; GABRP; GABRQ; GABRR1; GABRR2; GABRR3; GLS2; GOT1; GOT2; CTPS2; SLC38A1; GLUL; SLC1A3; SLC1A2; SLC1A1; SLC1A6; SLC1A7; SLC1A4; SLC17A6; SLC17A7; SLC17A8; GRIA1; GRIA2; GRIA3; GRIA4; GRIK1; GRIK2; GRIK3; GRIK4; GRIK5; GRIN1; GRIN2A; GRIN2B; GRIN2C; GRIN2D; GRIN3A; GRIN3B; GRM1; GRM2; GRM3; GRM4; GRM5; GRM6; GRM7; GRM8; SHMT1; SHMT2; GCSH; GLDC; AMT; DLD; SLC6A9; SLC6A5; SLC38A5; GLRA1; GLRA2; GLRA3; GLRA4; GLRB; HDC; HNMT; ALDH2; HAL; DAO; HRH1; HRH2; HRH3; HRH4; DBH; MAOA; MAOB; ADRA1A; ADRA1B; ADRA1D; ADRA2A; ADRA2B; ADRA2C; ADRB1; ADRB2; ADRB3; TPH1; TPH2; ALDH1A1; SLC6A4; SLC29A4; HTR1A; HTR1B; HTR1D; HTR1F; HTR2A; HTR2B; HTR2C; HTR3A; HTR3B; HTR3C; HTR3D; HTR3E; HTR4; HTR5A; HTR6; HTR7; MTNR1A; MTNR1B; ASMT; AANAT | 5HT1a; 5HT1b; 5HT2a; 5HT4; 5HT6; 5HTT; A4B2; A4B2; CB1; D1; D2; DAT; GABAa; GABAa; GABAa; H3; M1; mGluR5; MU; NAT; VAChT | HTR1A; HTR1B; HTR2A; HTR4; HTR6; SLC6A4; CHRNA4; CHRNB2; CNR1; DRD1; DRD2; SLC6A3; GABRA1; GABRB2; GABRG2; HRH3; CHRM1; GRM5; OPRM1; SLC6A2; SLC18A3 | TH; DDC; PAH; QDPR; TYR; DCT; SLC18A1; SLC18A2; CDNF; GCH1; SPR; TYRP1; CYP2D6; DRD1; DRD2; DRD3; DRD4; DRD5; DAT; MAOA; MAOB; COMT; SLC6A2; SLC6A3; DBH; PCBD1; PTS; CALY; PPP1R1B | LRRK2; SNCA; VPS35; PARK7; PRKN; PINK1; CHCHD2; DJ1; GBA; ATXN2; KIAA1840; ZFYVE26; POLG; DNAJC12; ATX; EPM2A; C9orf72; GRN; MAPT; PDE8B; PDGFRB; XPR1; ATP13A2; PLA2G6; DCTN1; DNAJC6; FBXO7; SYNJ1; VPS13C; RAB39B; JAM2; SLC20A2; ATP1A3; TAF1; GCH1; TH; SPR; QDPR; PTS; SLC6A3; SLC30A10; GLB1; WDR45; CP; FTL; FA2H; C19orf12; PANK2 | TOR1A; THAP1; GNAL; ANO3; EIF2AK2; HPCA; KMT2B; VPS16; PRKRA; GCH1; TH; ATP1A3; TAF1; SGCE; ADCY5; HPRT; ACAT1; GCDH; MUT; PCCA; PCCB; DCAF17; DDC; SLC30A10; SPR; QDPR; PTS; SLC6A3; PANK2; PLA2G6; ATP7B; SLC19A3; TIMM8A; ND6; GLB1; CP; SUCLA2; TUBB4A; ATXN3; WDR45; FTL; FA2H; KIF1C; C19orf12 | LRRK2; SNCA; VPS35; PARK7; PRKN; PINK1; CHCHD2; DJ1; GBA | TOR1A; THAP1; GNAL; ANO3; EIF2AK2; HPCA; KMT2B SGCE TAF1 TUBB4A AOPEP | WDR6; ARIH2OS; C3orf84; CCDC71; CTNND1; DALRD3; EFNA1; HCN1; IP6K2; MED19; NCKIPSD; NDUFAF3; P4HTM; PRKAR2A; PSMG1; SFMBT1; STIMATE; THBS3; TMX2; UNC5D; USP19 |

**Table S2:** Datasets and references of lesion network mapping studies
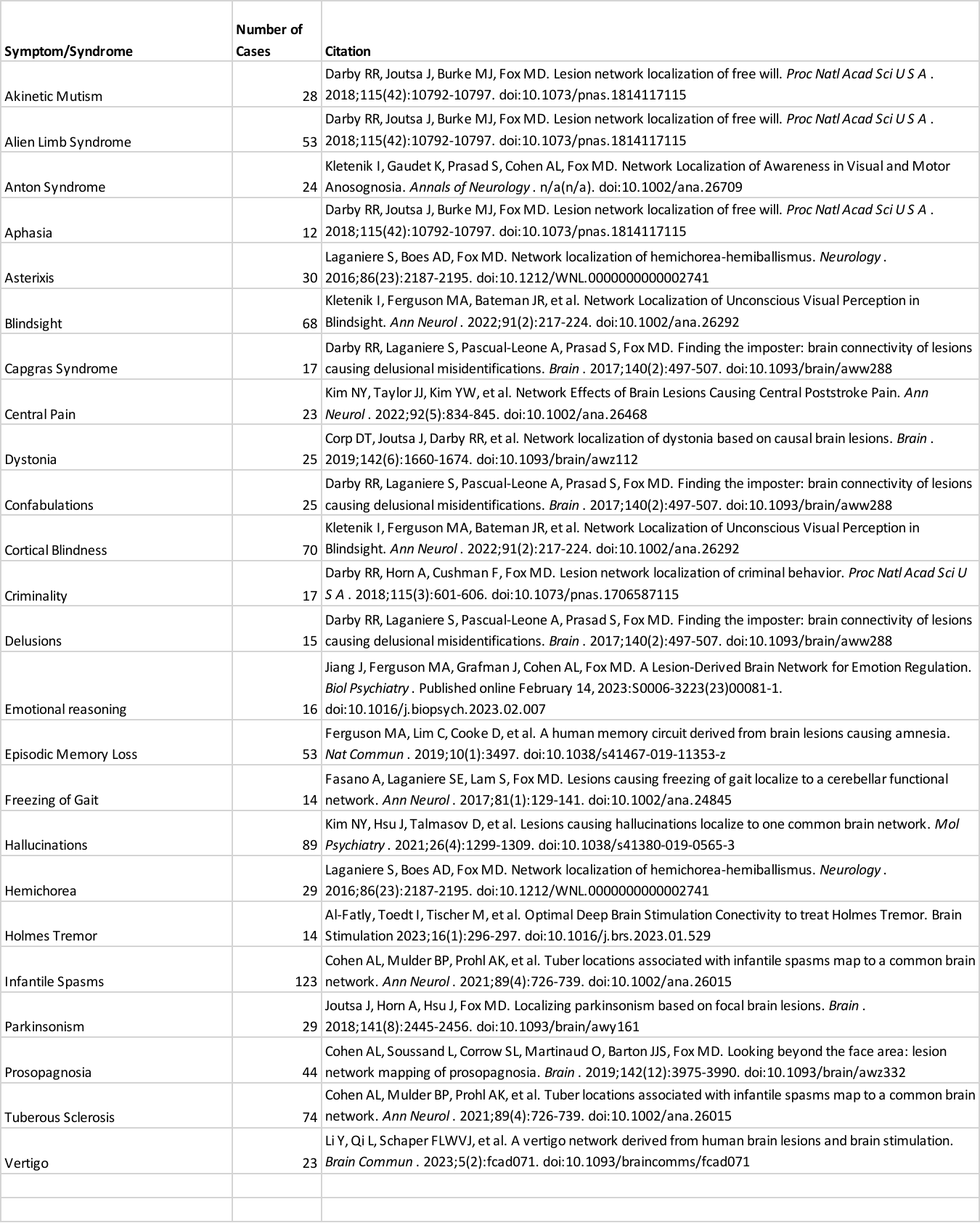
used as control groups in the Harvard Lesion Repository.

**Table S3:** Description and references of DBS datasets used for connectivity analysis.

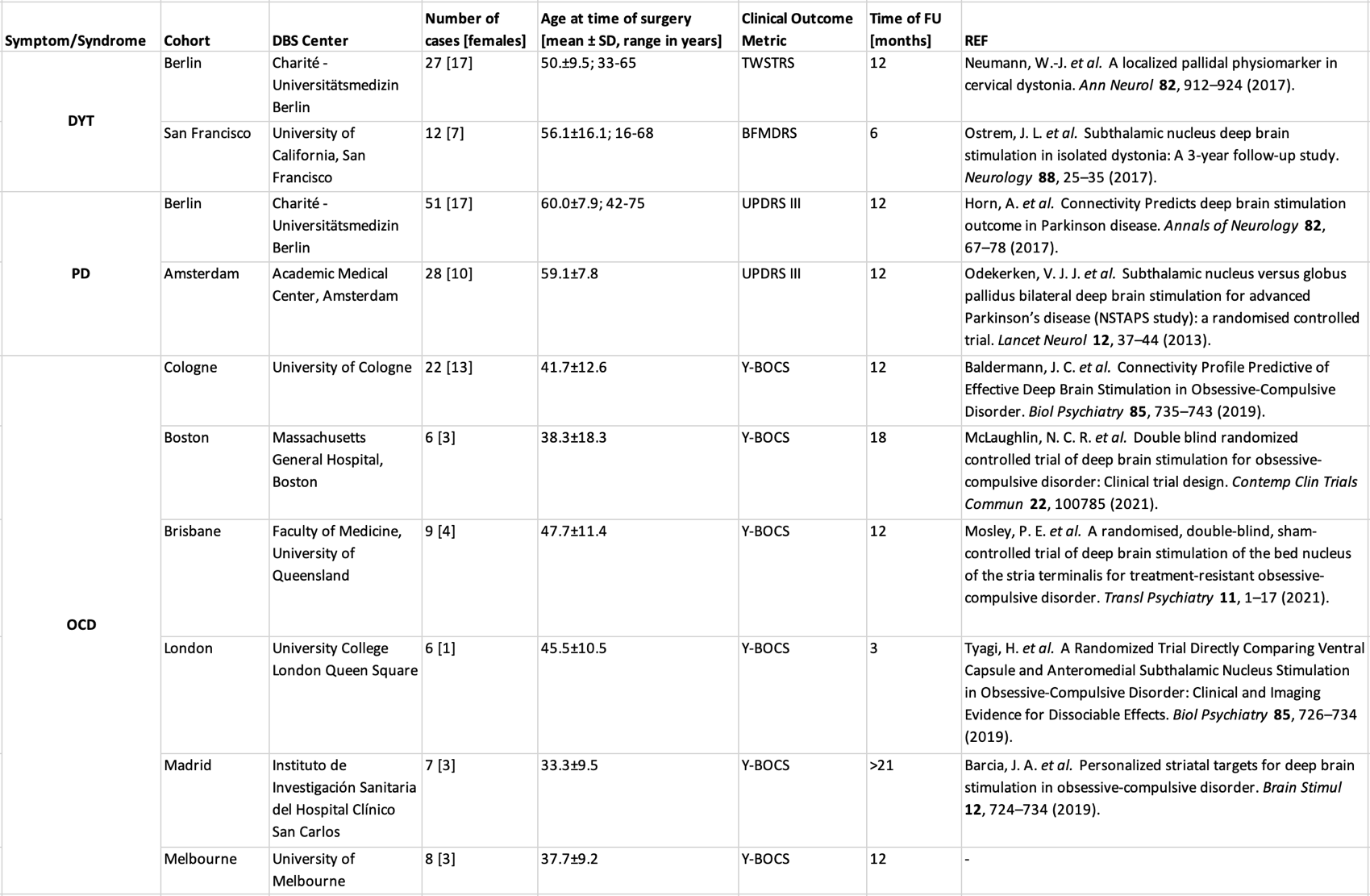

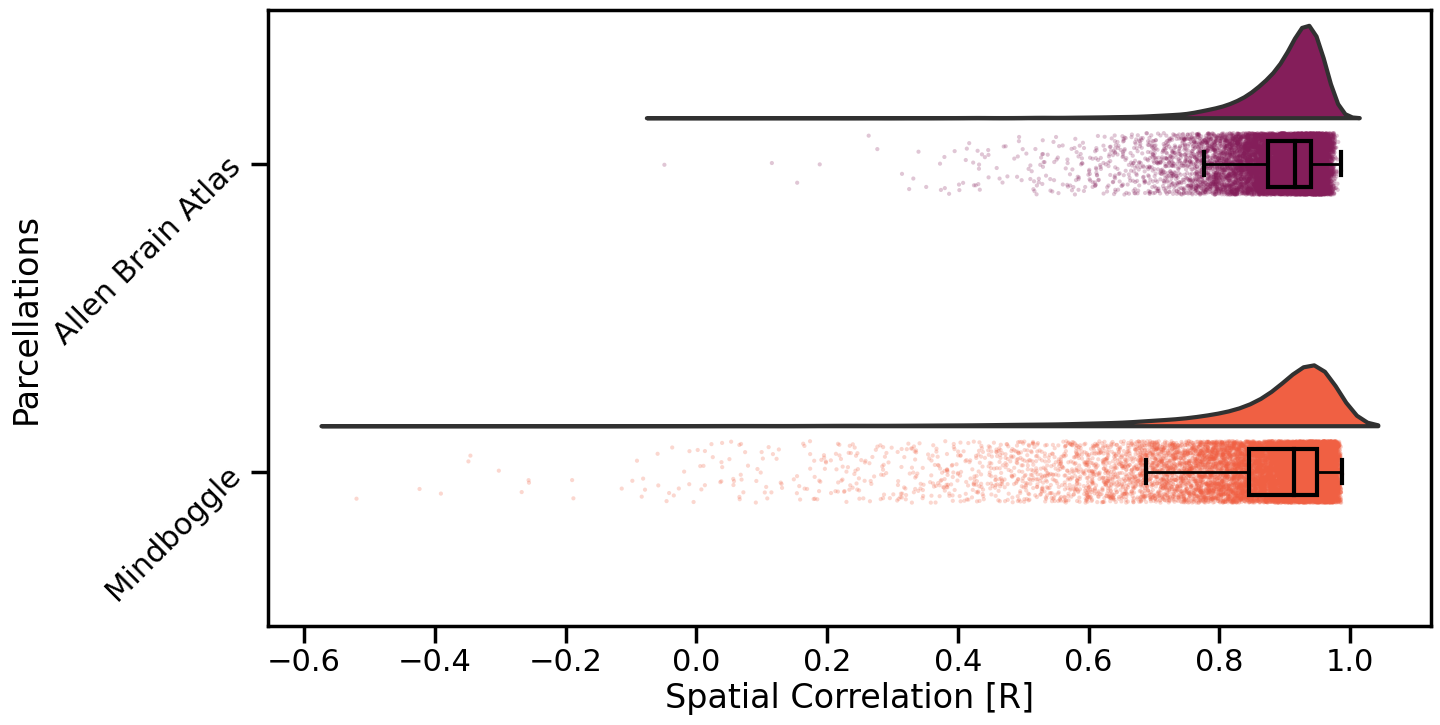

**Fig. S1: Comparison of different atlas parcellation on spatial variability in generated gene network maps.** Spatial correlation of gene network maps generated using a 430-region whole-brain volumetric atlas and gene network maps generated using parcellations from the Allen Human Brain Atlas(*95*) and the Mindboggle 101 Atlas(*104*), respectively. Raincloud plots feature the distribution of spatial correlations between pairs of all 15,632 gene network maps based on different parcellation schemes (average R across all genes comparing the 430-region atlas to the Allen Brain Atlas: 0.89±0.07 [mean±SD]; average R across all genes comparing the 430-region atlas to the Mindboggle Atlas: 0.87±0.14 [mean±SD]).

**Validation of Gene-Network Maps Using Pharmacological MRI**

Gene-network maps were validated by comparing them to brain function as measured by pharmacological MRI (phMRI). Specifically, we investigated whether gene network maps can explain variance in functional connectivity during pharmacologically induced perturbations.

Pharmacotherapy leverages the existing neurotransmitter receptor and transporter landscape to functionally alter brain states by targeting individual, or multiple receptors that show affinity for the specific drug(*37*). These changes can be captured by pharmacological MRI (phMRI), where individuals are scanned under the influence of a given pharmacological agent. As a result, phMRI identifies the functional consequences of neurotransmitter receptor engagement, providing a window into the brain’s altered network activity during pharmacological intervention.

Here, we leveraged a publicly available phMRI dataset comprising fMRI data acquired in 224 subjects under the influence of various pharmacological agents, including ayahuasca, lysergic acid diethylamide (LSD), 3,4-methylenedioxymethamphetamine (MDMA), N,N-Dimethyltryptamine (DMT), psilocybin, ketamine, methylphenidate, modafinil, and the anesthetics propofol and sevoflurane (Supplementary Table 1). In the dataset, functional connectivity was defined as the average (across subjects) of the within-subject difference in regional functional connectivity weighted degree between task-free fMRI scans at baseline and under the drug’s effects (see also Luppi et al.(*37*)). We hypothesized that gene network maps associated with neurotransmitter synthesis, regulation, and function would show similar connectivity patterns as maps of drug-induced functional connectivity reorganization.

To test this hypothesis, we calculated co-variance matrices reflecting pharmacological co-susceptibility and gene network co-activation between pairs of brain regions defined by an atlas parcellation used in the publicly available phMRI dataset (Supplementary Figure 2)(*37*). To generate co-variance matrices, we first computed a region-by-connectivity matrix for each modality, where each column represented either the functional changes induced by a specific pharmacological agent, or the region-wise gene-network activation for an individual gene. The rows in this matrix reflected the regional differences in connectivity across pharmacological agents and gene-network activation, respectively. Next, we performed pairwise correlations of the rows to generate co-variance matrices. These matrices quantified the relationships between pharmacological effects (pharmacological co-susceptibility) and gene network activations (gene-network co-activation) across brain regions, respectively. Finally, we compared the lower triangles of the pharmacological co-susceptibility and gene-network co-activation maps by correlating them to assess the similarity between drug-induced connectivity changes and gene-network activation patterns. This approach allowed us to test whether regions with similar drug-induced connectivity changes also exhibited similar gene-network activations.

Supporting our hypothesis, we found that *gene-network co-activation* is significantly correlated with pharmacological co-susceptibility (R=0.46, p<0.001/ ρ=0.46, p<0.001). This suggests that gene-network maps related to neurotransmitter synthesis, regulation, and function, reliably capture connectivity patterns induced by pharmacological agents. To ensure that this finding was not solely driven by shared variance in the connectome, we repeated this analysis, using partial correlation, controlling for raw connectivity. Even after accounting for this factor, the analysis explained a significant amount of variance (R=0.22, p<0.001; ρ=0.22, p<0.001).

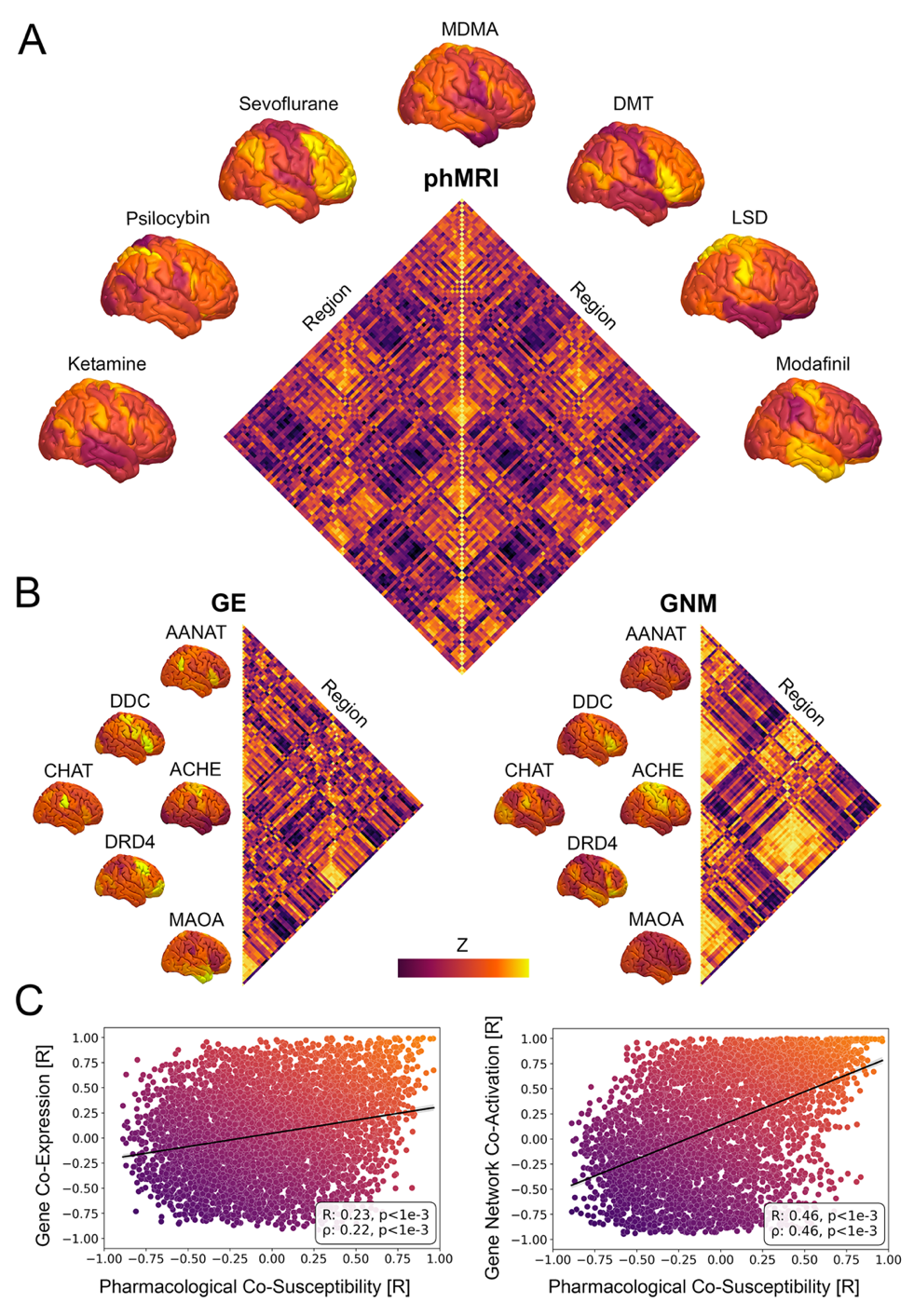

**Fig. S2: Relationship of local and network chemoarchitecture with phMRI responses. (A)** A region-by-region matrix of pharmacological co-susceptibility was generated by pairwise correlation of patterns of drug-induced functional connectivity changes for different pharmacological agents. Example phMRI maps featuring pharmacologically induced functional network reorganization patterns are displayed (see Supplementary Table 1 for a full description of the dataset) **(B)** Region-by-region matrices of local gene co-expression (GE, left) and gene network co-activation (GNM, right) were obtained in a similar manner. Brain surfaces feature local expression and network patterns of example genes (see Supplementary Table 1 for a full description of the dataset). **(C)** Correlation of phMRI and GNM matrices explained more variance compared to correlation of phMRI and GE matrices, capturing pharmacotherapy-induced connectivity changes more effectively than local gene expression.

**Validation of Gene-Network Maps Using PET**

Next, we sought to determine whether maps of gene-network co-activation offer advantages over maps of *local* gene expression (i.e., raw transcriptomic maps without information added by the functional connectome). To investigate this, we used the same neurotransmitter genes related to synthesis, regulation, and function as described earlier (Supplementary Table 1) to build neurotransmitter gene co-expression matrices, reflecting the co-variance of *local* neurotransmitter gene expression patterns across brain regions. As expected, neurotransmitter gene *co-expression* matrices significantly correlated with pharmacological co-susceptibilities but could only explain 5% of the observed variance (R=0.23, p<0.001/ ρ=0.22, p<0.001; controlling for raw connectivity: R=0.06, p<0.001/ ρ=0.06, p<0.001), as compared to 21% of the variance explained by gene-network maps. In other words, while both gene expression and gene network maps were able to capture regional variance in pharmacotherapy-induced functional connectivity changes, significantly more variance was explained on a gene *network* level (Z=15.01, p<0.001).

In a final validation step, we compared our findings to openly available positron emission tomography (PET) data(*36*). Given that PET measures neurotransmitter receptor densities at the local level, we hypothesized that local gene co-expression matrices would more closely resemble PET co-localization maps as compared to gene-network co-activation matrices. Results are detailed in Supplementary Figure 3. In brief, as expected, PET colocalizations were able to explain more variance in gene co-expressions (R=0.51, p<0.001/ ρ=0.49, p<0.001; Supplementary Figure 3C) as compared to gene-network co-activation (R=0.35, p<0.001/ ρ=0.34, p<0.001; Supplementary Figure 3C) highlighting that regional quantification of molecular markers and their expression may not sufficiently associate with the network dynamics observed during phMRI.

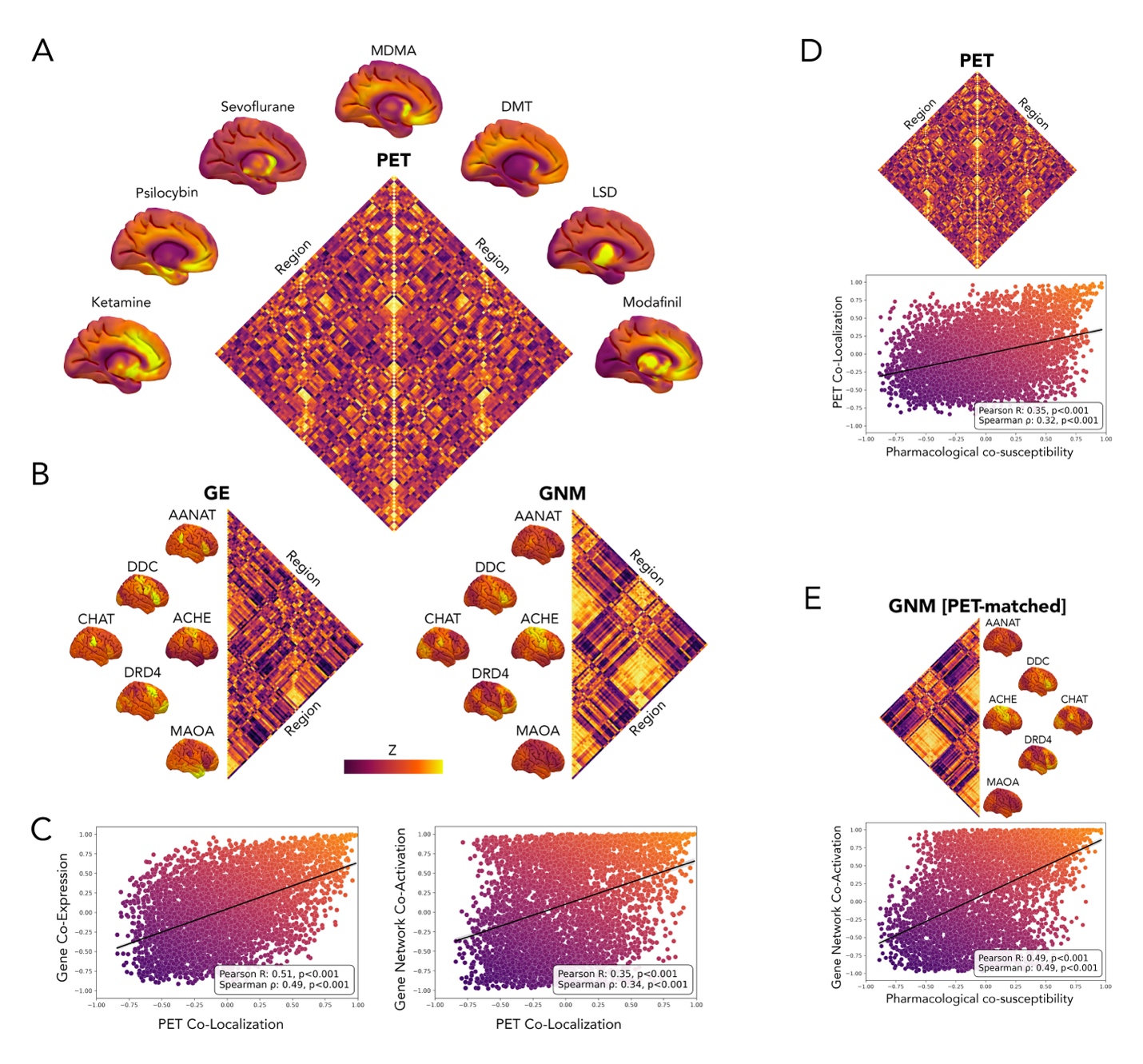
 **Fig. S3: PET Control Analyses.** (A-C) Correlation of covariance matrices associated with PET Co-Localization vs. gene co-expression and gene network co-activation. PET co-localization was able to explain more variance in gene co-expression (R=0.51, p<0.001/ ρ=0.49, p<0.001) as compared to pharmacological co-susceptibility (R=0.35, p<0.001/ ρ=0.34, p<0.001). (D) Correlation of pharmacological co-susceptibility and PET Co-Localization (E) Correlation of pharmacological co-susceptibility with gene network co-activation using only genes coding for neurotransmitter receptors identified through PET imaging.
